# PREDICTIVE VALUE OF SMELL AND TASTE TEST VS PCR-RT SARS-COV-2 AND RAPID DIAGNOSTIC TESTS IN THE DIAGNOSIS OF INFECTION BY COVID-19. A PROSPECTIVE MULTI-CENTRIC STUDY

**DOI:** 10.1101/2020.08.31.20185298

**Authors:** Pieruzzini Rosalinda, Ayala-Grosso Carlos, Navas José de Jesús, Rodríguez Wilneg Carolina, Parra Nathalia, Luque Emily, Sánchez Gago Aida, González Scarleth, Hagobian Alexandra, Grullón Angeline, Díaz Karen, Morales Mariano, De Jesús Melanie, Peña Sonia, Rodríguez Luis, Peña Luis Lenin, Asaro Ana, Magris Magda

## Abstract

There is a relationship between smell and taste disturbances and coronavirus infection. These symptoms have been considered the best predictor of coronavirus infection, for this reason, it was decided to evaluate the predictive value of the smell and taste test and its association with the results of SARS-CoV-2 PCR-RT and rapid diagnostic tests. in the diagnosis of pathology. Methodology: 248 patients divided into 3 groups: asymptomatic, symptomatic without chemosensory disorders, and chemosensory disorders alone. All of them underwent SARS-CoV-2 PCR-RT, a rapid diagnostic test and a test of Venezuelan smell and basic taste at the beginning. Weekly follow-up with smell and taste test and SARS-CoV-2 PCR-RT until recovery. Results: 20.56% of patients had smell and taste disorders to a variable degree and were positive by SARS-CoV-PCR-RT. 2.15.3% of patients with chemosensory disorders were negative for COVID-19. The positive predictive value of the smell and taste test was 57.3; Sensitivity 41.13% and specificity 69.35%. There were no statistically significant differences by age, sex and chemosensory disorders. The predominant chemosensory disorder was the combination of mild hyposmia and hypogeusia and appeared in the company of other symptoms. Recovery occurred in an average of 8.5 days, asynchronously with the SARS-CoV-2 RT-PCR negativization, which occurred up to more than 15 days after the senses recovered. Maximum time of negativization of the RT-PCR of 34 days. Conclusion: chemosensory disorders are a symptom and / or sign of coronavirus disease but cannot be considered as predictors of said disease in this population studied. The gold standard remains the SARS-CoV-2 PCR-RT test. Rapid diagnostic tests should be used for follow-up. Recommendations: it is necessary to expand the sample, include routine psychophysical smell and taste tests to screen cases and take race and virus mutations into consideration to explain behavior in certain populations.

## INTRODUCTION

The incidence of smell and taste disorder in the early phase of SARS-CoV-2 infection appears to be of predictive value in the course of the infection. Since the beginning of the pandemic, multiple studies have reported a highly variable association between olfactory dysfunction and the presence of other characteristic symptoms of coronavirus infection. The first European multicenter study, which collects clinical data from 12 hospitals, established that in an examined population of 417 patients with mild to moderate presentation of COVID-19, the most common symptoms were cough, myalgia and loss of appetite; while the symptoms related to otorhinolaryngology disorders were facial pain and nasal obstruction. At least 86% of the cases had olfactory alterations and 88% had taste alterations. Olfactory dysfunction appeared before another symptom in 11.8% of cases (1)

The observation of a sudden loss of smell in association with the increase in cases of coronavirus was reported in the United Kingdom, recently, however, the evidence seems to be circumstantial since it was found as information provided in the social networks of patients who presented this symptom in isolation. The authors reported 9 cases of sudden anosmia without other associated symptoms in the first 3 weeks of the onset of the coronavirus, and it should be noted that none of these patients was evaluated with a specific test to assess the presence of the disorder. (2)

More recently, in another study carried out at the L. Sacco Hospital in Milan, Italy, it is established as a result of a survey of 59 patients out of 88 admitted that 33.9% had at least one smell and / or taste disorder and 18.6 % both. Of this group of patients, 20.3% presented olfactory symptoms prior to their admission to the hospital and 13.5% during the stay. (3)

The frequency of olfactory alterations associated with a viral infection is not new in otorhinolaryngology; many viruses can cause olfactory dysfunction due to an inflammatory process in the nasal mucosa and the development of rhinorrhea. Among the viral agents that are associated with these alterations are rhinovirus, parainfluenza, Epstein-Bar and some coronaviruses (4,5).

However, the observation that the olfactory dysfunction associated with SARS-CoV-2 is not essentially associated with the appearance of rhinorrhea and nasal obstruction suggests a different mechanism of action. Until now, the pathophysiology of taste and smell disorders in SARS-CoV-2 infection is unknown. One of the most relevant evidences on the mechanism of action of the coronavirus has been presented in an investigation carried out in the olfactory mucosa of mice and analyzing data from human RNA sequences, in which it was found that the 2 genes express the information for the ACE2 and TMPRSS2 receptors involved in the entry of COV-2 into the cell, are expressed in the cells of the respiratory epithelium of the nasal cavity, support cells, Bowman’s glands, microvilli and stem cells of the olfactory mucosa, but not in olfactory sensory neurons, suggesting that the olfactory damage mechanism is non-neural. Regarding damage to the sense of taste, it seems to be directly on the taste receptor and the production of cytokines that irritate the trigeminal and glossopharyngeal nerves related to the transmission of the nerve signal to the central nervous system. (6)

Netland et al, determined in some transgenic mice an extensive replication of the virus in the brain, with some regional differences and that this brain infection, in turn, was an important factor in the aspiration pneumonia observed in some of the mice studied. The generalized neuronal infection occurred after intranasal inoculation, which suggests that its entry point may be through the olfactory bulb. This raises questions about the possible future effects of the coronavirus in patients suffering from SARS-CoV-2 infection (7).

However, the mechanisms by which taste and olfactory tissue damage occurs during SARS-CoV-2 infection remain unclear.

A large majority of initial reports on alterations in smell and taste in relation to coronavirus infection were based on results collected by patient surveys, without corroborating the diagnosis by psychophysical tests of chemosensory alterations. Patients were not followed up systematically until recovery, nor was an association established with diagnostic tests such as SARS-CoV-2 RT-PCR or rapid diagnostic tests. These factors have been able to influence the overestimation of the symptom as a predictor of the disease. Similarly, the use of olfactory tests not adapted to the population studied may lead to the presence of false positives and consequently an overestimation of the diagnosis of chemosensory alterations.

The previously described has motivated us to carry out an investigation, whose distinctive character is the determination of the predictive value of the smell and taste tests in the diagnosis of SARS-CoV-2 infection and its relationship with other diagnostic tests such as CRP -RT SARS-CoV-2 and rapid diagnostic tests as well as monitoring and comprehensive observation of the patient’s recovery from coronavirus infection.

In Venezuela, the first case of COVID-19 was reported in March of this year with a linear behavior and a low number of cases reported until May, in which patients increased exponentially and the severity of the clinical presentation has varied in some foci of the country in relation to others, so that the results obtained may change due to the penetration of the virus in the population and some predominant migratory characteristics in the country.

## METHODOLOGY

### POPULATION AND SAMPLE

During the massive screening for SARS-CoV-2 infection, 500 suspected cases of COVID-19 were evaluated, from the Capital District and the states of Miranda, La Guaira and Nueva Esparta, which presented the highest concentration of positive cases while it was carried ou this investigation. All individuals were: a. SARS-CoV-2 Rapid Diagnostic Test; b. Test of the Venezuelan smell and basic taste. Of these, the individuals who gave their informed consent to participate in the study were selected and excluded: a. patients with acute or chronic rhinosinusitis, allergic rhinitis, obstructive rhinopathy previously known to the patient; b. patients with a history of exposure to toxic, irritating chemicals that cause alterations in the smell and taste and; c. patients with diabetic neuropathy of more than 1 year of evolution.

Of the 248 individuals who met the selection criteria, they were divided into three groups (Table 1) and the following were performed: 1. Complete medical history that included age, sex, epidemiological history of travel and contacts with positive cases, questioning of suggestive respiratory symptoms and other symptoms, presence or absence of smell and / or taste disorders. 2. General ENT physical examination, Venezuelan smell test and basic taste test. 3. Rapid Diagnostic Tests for SARS-CoV-2 and nasopharyngeal sample collection for SARS-CoV-2 RT-PCR at the time of protocol entry. Then, weekly follow-up of SARS-CoV-2 RT-PCR tests and psychophysical tests were carried out until recovery of the chemosensory disorder and negative RT-PCR. (Table 1)

**Table no. 1.** Distribution of study patients

| Subjects | n (%) | Description |
| --- | --- | --- |
| Asymptomatic | 104(41,93%) | Patient who previously did not present symptoms or signs related to the coronavirus |
| Symptomatic | 55(22,17%) | Patients who presented some of the following symptoms: headache, cough, fever, general malaise, chest pain, dyspnea, myalgia, arthralgia, chills, hyporexia, nausea, vomiting, diarrhea and any symptoms that have been described in the literature that were present and / or was suggestive of SARS-Cov-2 infection, not involving smell and / or taste disorders. |
| Smell and taste disorders | 89(35,38%) | They presented some alteration of smell and / or taste, determined by the corresponding psychophysical tests. |
| Total | 248(100%) |  |

## CLINICAL ASSESSMENT

### Smell and Taste Test

The smell test used is a short adaptation of the Test of the University of Pennsylvania (UPSIT), which consists of 2 notebooks with 10 odorant substances (5 in each, for a total of 10) typical of the Venezuelan that were obtained from a study made in more than 1000 people. The substances are: coffee, chocolate, baby cologne, scented talcum powder, liquid detergent, coconut essence, powder detergent, cinnamon, acetone and rum (8,9). The patient must identify the corresponding odor on each page of the booklet and distinguish between the distractors that are specified. It was decided to use an adapted and previously validated test for the Venezuelan population, because some of the odorant substances from other psychophysical tests were not recognized by the patients and were susceptible to diagnostic errors. Furthermore, the Venezuelan test is inexpensive, easy to make, and can be applied on a large scale. The quantitative scale for the diagnosis of olfactory alterations with the Venezuelan test is dependent on the number of recognized odorants:

Normosmia: 8-10

Mild hyposmia: 6-7

Moderate hyposmia: 5-4

Severe hyposmia: 2-3

Anosmia: 0-1

The taste test was performed with the 5 basic flavors universally described: sweet, salty, acid, bitter and umami (10) and the patient was diagnosed with ageusia if he did not recognize flavors and hypogeusia if the patient recognized up to 4 flavors.

There is a qualitative diagnostic category for both olfactory and taste alterations, but they were not evaluated in this study.

## TESTS FOR SARS-CoV-2 RAPID DIAGNOSIS (PDR)

Diagnostic tests No. IFU-COVID3-01 were used. Brand ANHUI DEEPBLUE MEDICAL TECHNOLOGY CO. LTD. Lot: 20200307 IgM-IgG.

It is an antibody test kit (Colloidal Gold) that is used for the qualitative detection of antibodies against the new coronavirus IgG / IgM in human serum, plasma and whole blood. The test uses the principle of the antigen of the new recombinant coronavirus (COVID-19) labeled with colloidal gold. A nitrocellulose membrane (NC) is sensitized with rat anti-human IgM and IgG antibodies and polyclonal sheep anti-rat antibodies; When the sample contains IgM antibodies, it comes into contact with the membrane (NC), a compound is produced with the coronavirus antigen labeled with colloidal gold, it is captured by the rat anti-human IgM antibodies, resulting in a colored line. If the sample contains IgG antibodies, it forms a compound with the antigen labeled with colloidal gold, this compound is captured by the rat anti-human IgG antibody, also resulting in a colored line. When the samples contain IgM and IgG at the same time, they form 2 lines. When there are no IgM or IgG antibodies in the sample, only one quality control line is marked and the result is negative.

These tests were performed for screening at the beginning of the study, however, patients were not excluded just because they could present a negative result. On the contrary, it was decided to make a correlation between the patients with the rapid tests, RT-PCR and psychophysical tests of smell and taste to determine their usefulness in the diagnosis and subsequent follow-up.

### COLLECTION AND TRANSFER OF RT-PCR SAMPLES for SARS-CoV-2

Patient samples were obtained with strict biosafety protocol and nasopharyngeal swab. They were placed in YOCON brand viral transport medium, lot Y25200101 and stored at a temperature between 2-8 C until the reference laboratory.

The SARS-CoV-2 PCR-RTs were processed and reported by the Rafael Rangel National Hygiene Institute of the Ministry of Popular Power for Health.

### ETHICAL APPROVAL

This research was reviewed and approved by the Bioethics Committee of the “Hospital University” Dr. Carlos Arvelo “, in addition each individual was informed about the study and voluntarily signed the informed consent.

**Figure nº1.**
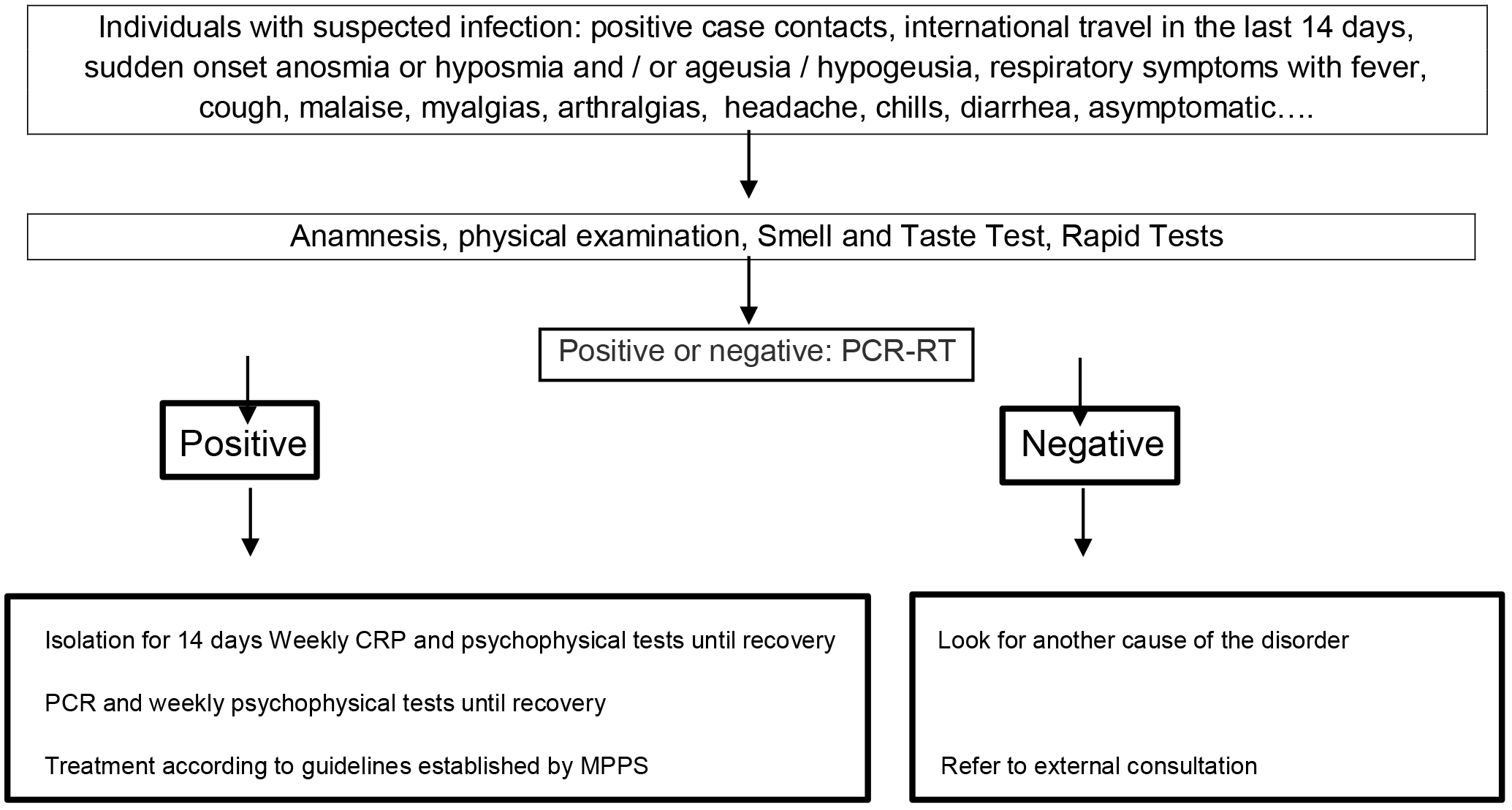
METHODOLOGICAL SCHEME

### STATISTIC ANALYSIS

Descriptive statistics and measures of central tendency were used for the analysis for the age groups and type of chemosensory disorder. In calculating the Predictive Value, Sensitivity and Specificity of the diagnostic tests used, the Wilson points method was used through the Openepi program, version 3. Open source calculator Diagnostic Test.

Regarding the correlations and statistical significance tests of the groups by gender, sex, and diagnosis of chemosensory disorders, the SPSS v20 program was used.

## RESULTS

Of a total of 500 patients in whom an initial screening was performed with rapid tests and chemosensory tests, 248 subjects were chosen. It is important to note that it was decided to reduce the study population, because it was observed that in the first 252 individuals, rapid tests were negative in 100% of cases, despite showing suspicious symptoms in many of them, which it included smell and taste disorders. These initial patients could not recover to complete the evaluation with RT-PCR for SARS-CoV-2 and their follow-up was difficult because some of them lived in areas with difficult access or in the interior of the country. Of the 248 subjects who completed the study, the following characteristics were observed:

**Table nºNo. 2.** Age and Sex of patients evaluated

|  | **M**<br>**n (%)** | **F**<br>**n (%)** | **Total**<br>**n (%)** |
| --- | --- | --- | --- |
| 18-24 años | 25(10,08%) | 13(5,24%) | 38 (15,32%) |
| 25-31 años | 39(15,72%) | 30(12,09%) | 69(27,82%) |
| 32-38 años | 29(11,68%) | 27(10,88%) | 56(22,58%) |
| 39-45 años | 13(5,24%) | 17(6,85%) | 30(12,09%) |
| 46-52 años | 16(6,45%) | 4(1,61%) | 20(8,06%) |
| 53-59 años | 16(6,45%) | 7(2,82%) | 23(9,27%) |
| 60-66 años | 3(1,20%) | 9(3,62%) | 12(4,83%) |
| Total | 141(56,85%) | 107(43,14%) | 248(100%) |

The predominant age groups were 25-38 years (50.4%) and males with 56.85%. Age average of 35.86 years.

**Table nº2.**
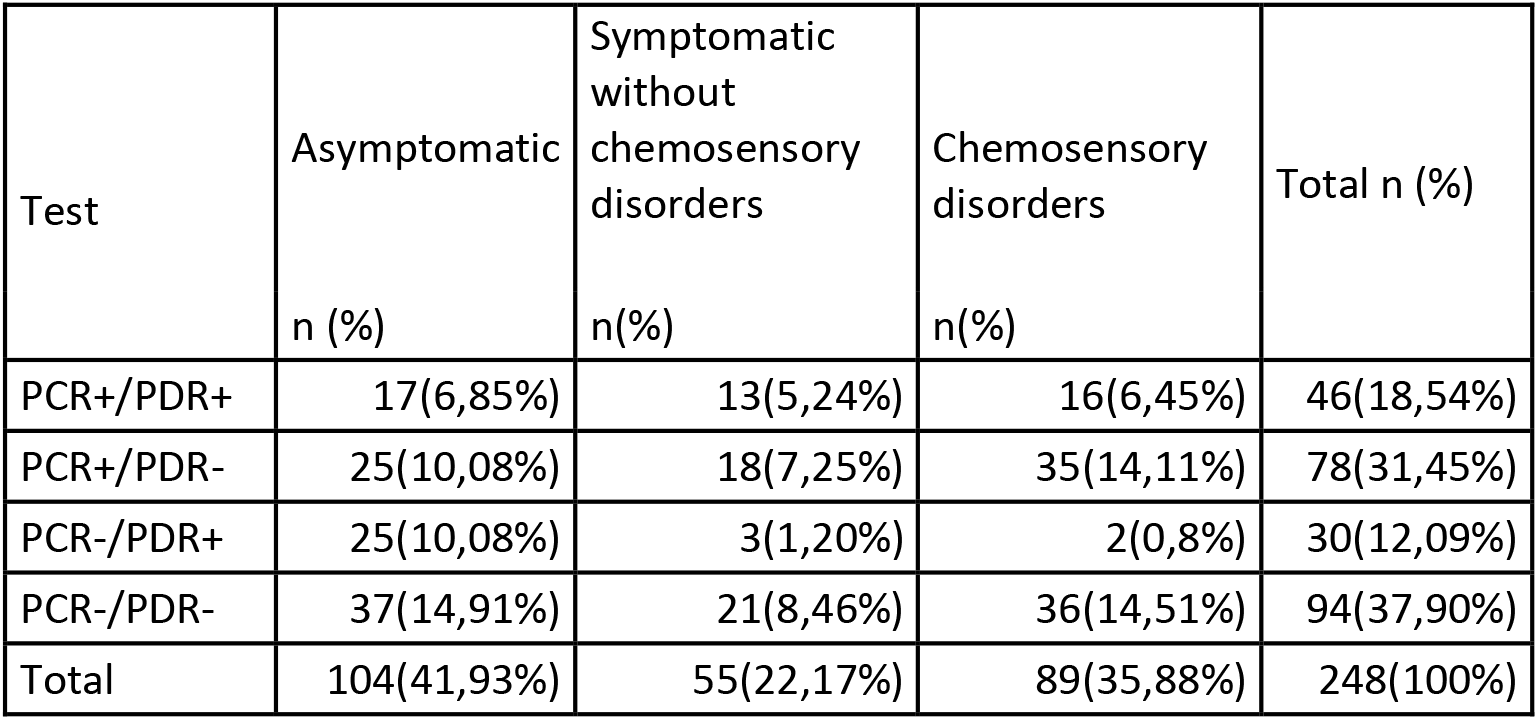
SARS-CoV-2 PCR-RT tests, rapid diagnostic tests (PDR), general symptoms and chemosensory disorders upon entering the protocol.

Of the 248 individuals evaluated with smell and taste tests, RT-PCR for SARS-CoV-2 and PDR IgM / IgG, 124 (50%) were SARS-CoV-2 PCR-RT positive, 42 (33.87%) of them were asymptomatic, 51 (41.12%) patients had smell and / or taste disorders or both verified by the test. The other 31 (25%) positive patients were general symptomatic. Note that there are 38 (15.32%) of the total cases that have smell and taste disorders and are negative by SARS-CoV-2 RT-PCR. 24 (9.6%) of negative symptomatic patients are also observed.

Regarding the results of the PDR tests, it can be seen that of the 248 cases, 172 were negative rapid test. This equates to 69.35% of the sample and only 30.64% (76) were SARS-CoV-2 PDR positive. From this, it is important to note that in 30 (12.9%) patients who were positive by rapid test, the definitive diagnosis by RT-PCR confirmed that they were negative for SARS-CoV-2. Reverse situation in 78 patients (31.45%) in whom the rapid test was negative and who finally were PCR-RT positive for SARS-CoV-2. This shows a limited usefulness of rapid tests for the initial diagnosis of SARS-CoV-2 infection.

**Table nº3.** Presence or absence of symptoms and SARS-CoV-2 infection.

|  | Asymptomatic<br>N(%) | Symptomatic<br>N(%) | Chemosensory disorders<br>N (%) | Total N (%) |
| --- | --- | --- | --- | --- |
| PCR+ | 42(16,93%) | 31(12,5%) | 51(20,56%) | 124(50%) |
| PCR- | 62(25%) | 24(9,67%) | 38(15,32%) | 124(50%) |
| total | 104(41,93%) | 55(22,17%) | 89(35,88%) | 248(100%) |

Table 3 shows that of the 248 individuals studied, 89 (35.88%) had taste and / or smell disorders, of these only 51 (20.56%) had SARS-Cov-2 infection by PCR -RT. Of the positive cases, 16.93% reported having no symptoms during the questioning, while 12.5% patients presented another symptom associated with COVID-19 that was not related to smell and / or taste disorders.

If only positive cases of SARS-CoV-2 infection are taken into account, which correspond to 124, then 41.12% (51) presented smell and taste disorders.

**Table No. 4.** Predictive Value of Chemosensory tests vs RT-PCR in the Diagnosis of Infection by SARS-Cov-2.

| Diagnostic Tests | PCR-RT positive | PCR-RT negative | Total |
| --- | --- | --- | --- |
| Test QS* positive | 51 | 38 | 89 |
| Test QS* negative | 73 | 86 | 159 |

\*QS test: Chemosensory test of smell and taste

The following values were obtained with a 95% confidence interval, using the Wilson points method: sensitivity of 41.13% and specificity of 69.35% of the chemosensory test in the diagnosis of COVID-19. The positive Predictive Value of the smell and taste test is 57.3% and the Negative Predictive Value is 54.09%. This means that a patient positive by RT-PCR for SARS-CoV-2 has 57.3% chances of having smell and taste disorders. The diagnostic accuracy of the chemosensory test versus the SARS-CoV-2 RT-PCR application is 55.24%.

**Table No. 5.** General symptoms and chemosensory disorders in SARS-CoV-2 infection

| Symptom type | PCR-RT+<br>n(%) |
| --- | --- |
| Chemosensory disorders alone | 15(18,29%) |
| Chemosensory disorders associated with other symptoms | 36(43,90%) |
| General symptoms | 31(37,80%) |
| Total | 82(100%) |

Only 15 (18.29%) of the patients reported the smell and taste disorder before carrying out the molecular diagnosis and before hospital admission as the only symptom and later corroborated by the psychophysical smell and taste tests (Table 5). To be detailed regarding the type of chemosensory alteration they presented, 5 (9.8%) had olfactory alterations alone, 2 only with anosmia, 9 (17.64%) combined alterations of smell and taste in variable degrees and 1 (1, 96%) taste alteration alone of the hypogeusia type.

43.90% (36) of the subjects, presented alterations of smell and taste in different degrees, associated with other symptoms such as fever (5), headache, myalgia, arthralgia, chills, odynophagia, hyporexia (20), dyspnea, dry cough and chest pain (2), myalgia alone (1), fever, headache, and general malaise (8). Of this group of symptomatic subjects, only 10 (27.77 %%) manifested chemosensory disorder before hospital admission. 26 patients (72.22%) required the chemosensory test to make the diagnosis of the alteration.

37.8% (31) SARS-CoV-2 RT-PCR positive individuals presented general symptoms without olfactory or taste disorders.

It should be noted that the analysis of this table has been carried out from the total number of patients with symptoms associated with COVID-19 (82/124 positive cases), excluding asymptomatic cases (42/124).

**Table nº6.** Age, sex and type of chemosensory disorder in SARS-CoV-2 patients.

| Female |  |  |  | Male |  |  |  |
| --- | --- | --- | --- | --- | --- | --- | --- |
| Type of disorder | 18-35 años<br>n(%) | 36-51 años<br>n(%) | > 52 años<br>n(%) | 18-35 años<br>n(%) | 36-51 años<br>n(%) | > de 52 años<br>N(%) | Total<br>N(%) |
| Anosmia | 0(0%) | 2(3,92%) | 1(1,96%) | 0 | 1(1,96%) | 1(1,96%) | 5 (9,8%) |
| Mild hyposmia | 5(9,8%) | 1(1,96%) | 1(1,96%) | 5(9,8%) | 4(7,84%) | 1(1,96%) | 17(33,33%) |
| Moderate hyposmia | 1(1,96%) | 0(0%) | 0(0%) | 1(1,96%) | 0(0%) | 1(1,96%) | 3(5,88%) |
| Severe hyposmia | 0(0%) | 0(0%) | 0(0%) | 1(1,96%) | 2(3,92%) | 0(0%) | 3(5,88%) |
| Hypogeusia | 3(5,88%) | 0(0%) | 1(1,96%) | 1(1,96%) | 0(0%) | 0(0%) | 5(9,8%) |
| Both of them | 3(5,88%) | 3(5,88%) | 1(1,96%) | 7(13,72%) | 1(1,96%) | 3(5,88%) | 18(35,29%) |
| Total | 12(23,52%) | 6(11,76%) | 4(7,84%) | 15(29,41%) | 8(15,68%) | 6(11,76%) | 51(100%) |

The presence of chemosensory disorders was observed in 29 male patients (56.86%) and 43.14% of cases (22) in female. This does not represent a statistically significant difference.

The most frequent chemosensory disorder was the combination of both disorders in 18 subjects (35.29%), followed by mild hyposmia in 17 patients (33.33%), hypogeusia in 9 patients (9.8%), moderate hyposmia in 3 cases (5.88%), severe hyposmia in 3 cases (5.88%)

The presence of both disorders in both sexes did not show statistically significant differences. Nor were significant differences by age observed. (p. <955 from 18 to 35 years old, p <.510 from 36 to 51 years old, p <.736 from over 52 years old) Table nº6

**Table No. 7.** Evolution of chemosensory disorders in patients with SARS-CoV-2 infection. Negative CRP-

| Olfactory disorder recovery time |  | PCR-RT- |
| --- | --- | --- |
| < 6 days | 19 | 0 |
| 7-14 days | 22 | 7 |
| 15-20 days | 6 | 19 |
| >21 days | 3 | 20 |
| No improvement (120days) | 1 | 0 |
| Undefined | 0 | 5 |

**Figure 2:**
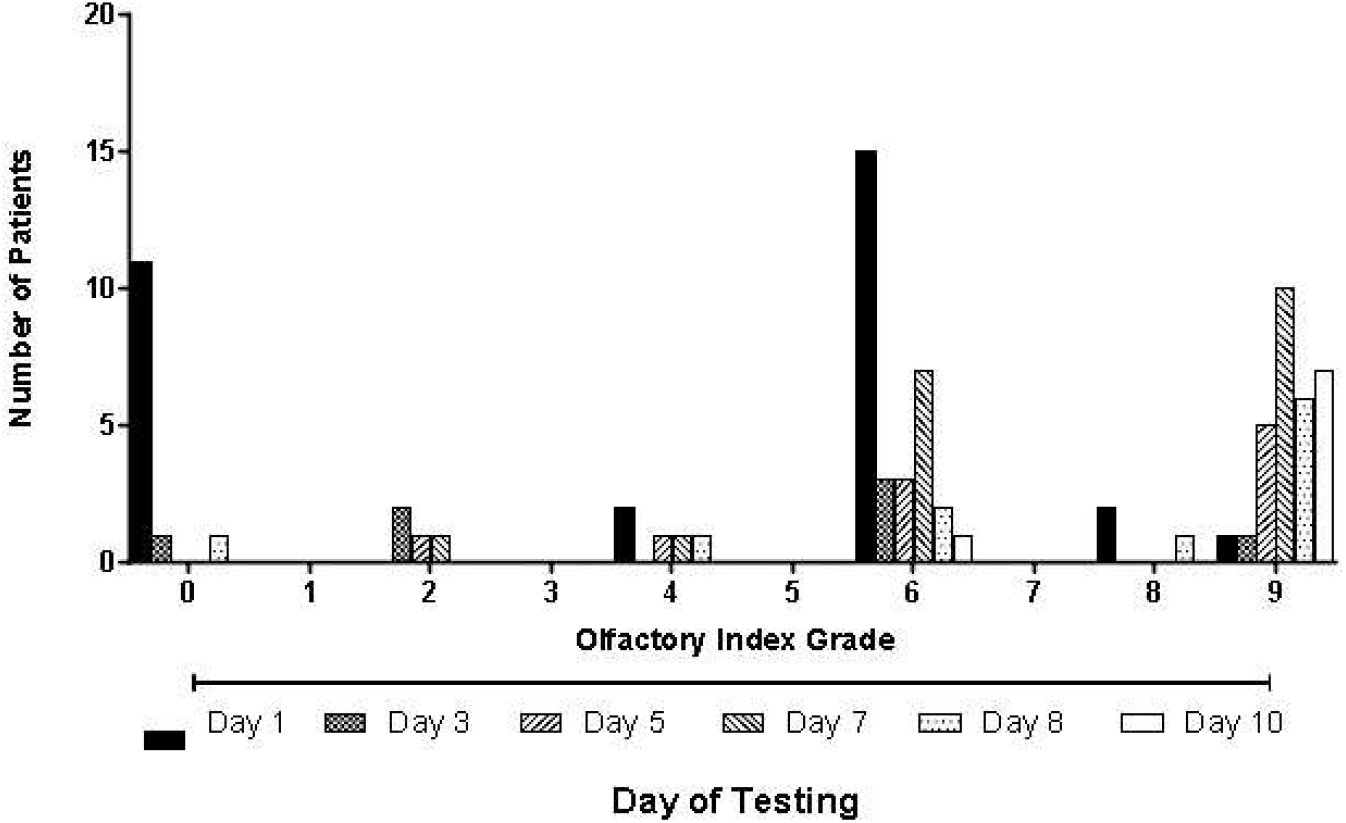
Olfactory index and average recovery from the disorder.In this image the data grouped by olfactory index are observed (mild hyposmia 6-7, moderate hyposmia 4-5, severe hyposmia 2-3, anosmia 0-1, normosmia 8-10) and days of recovery. The predominant type of olfactory disorder was mild hyposmia, with an average of 8.5 days recovery.

The average number of days of recovery from chemosensory disorder is 8.5 days and the RT-CRP becomes 76% negative in a period greater than 15 days after the improvement of the disorder. The patient with the shortest time of negative PCR was 7 days and the longest 34 days. There were 5 undefined cases, because they have persisted positive to date or the correct follow-up was not carried out. Average time to negative results by SARS-CoV-2 RT-PCR of 20 days. Maximum time of negativization of the RT-PCR of 34 days (table 7 and figure 3).

**Fig. 3.**
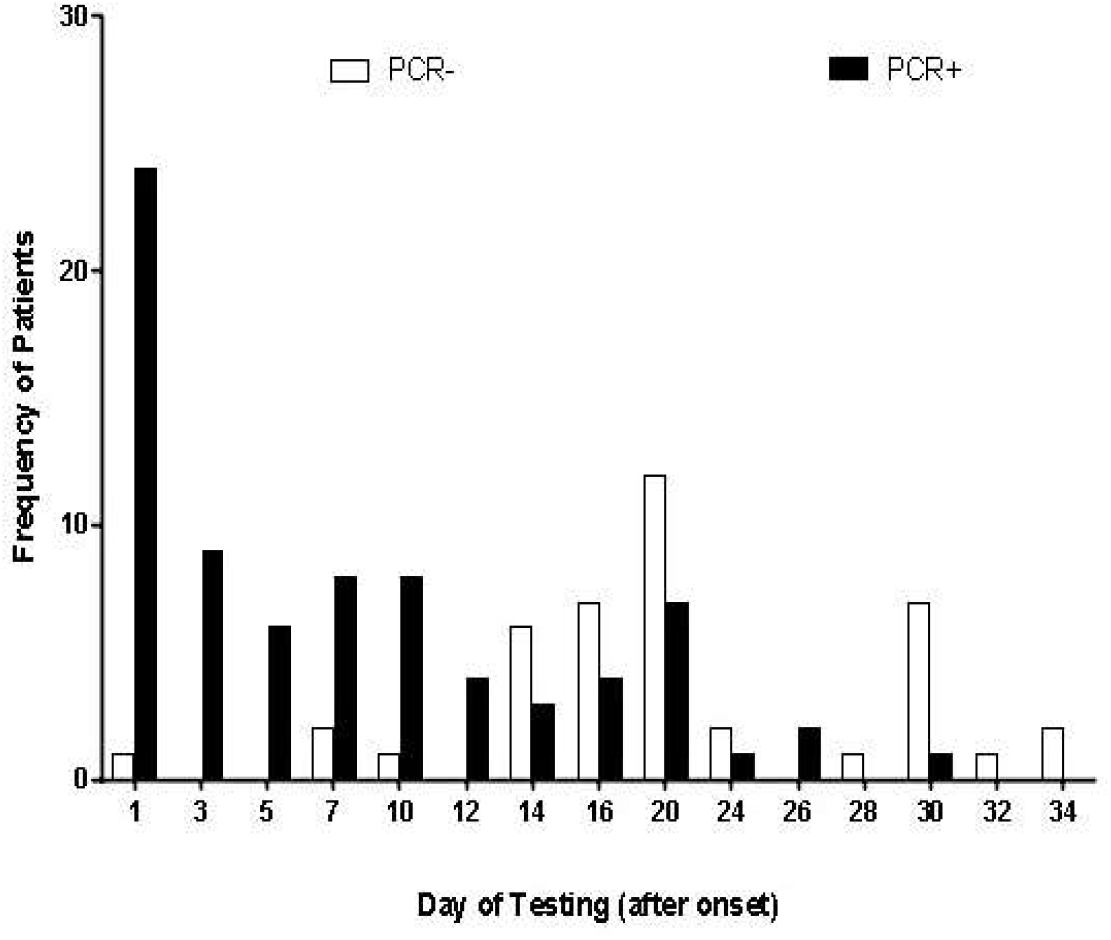
Progression of RT-PCR in patients with altered smell and taste

## Discussion

The objective of this research was to determine the predictive value of the smell and taste test in coronavirus infection and to characterize the symptoms and chemosensory disorders in the study population, both patients infected and not infected by SARS-CoV-2. A fundamental difference of this research in relation to other series, is that they were evaluated with smell and taste tests, rapid diagnostic tests and SARS-CoV-2 PCR-RT in 3 groups of patients, asymptomatic, general symptomatic and with disorders chemosensory disorders and from there we proceeded to determine their characteristics. We observed that chemosensory disorders are present during the coronavirus infection, but not as predictive factors of the disease as described in most series and may also be present in a significant number of patients without demonstrating a positive result for SARS-CoV-2 RT-PCR, which is a reason future research or further monitoring.

Series such as those published by Lechien et al (1) Gane et al (2) Spinato et al (11) revealed a high frequency of smell and taste alterations that ranged between 19.4 and 88% in patients with coronavirus infection In these studies, objective assessment of symptoms was not performed, but data were collected through surveys. It is important to note that targeting the symptom is a priority to corroborate the diagnosis, since there are patients who may perceive that they have a disorder but in fact, when examining it exhaustively, it is not. The opposite can also happen, the person is considered asymptomatic, but may carry a mild or qualitative disorder manifested by the inability to discriminate or confuse certain smells and / or flavors.

Vaira et al. (12) performed smell and taste tests in 72 positive patients at the University Hospital of Sassari, and 73.6% of the patients reported having chemosensory alterations. This work showed hyposmia to a variable degree in 60 cases and anosmia in 2 patients. The taste evaluation showed hypogeusia in 33 cases and complete ageusia in 1 patient.

Moein et al. (13) reported that 98% (59 of 60) of the patients had some type of olfactory loss using the UPSIT test. We included 248 patients divided into the 3 groups, as explained initially. We found that, of 124 positive cases for COVID-19, only 51 patients had smell and taste disorders, to detail 28 patients with smell disorders to varying degrees, 5 with anosmia, 17 with mild hyposmia, 3 with moderate hyposmia and 3 with severe hyposmia. 18 patients presented a combination of smell and taste disorders (7 with anosmia, 2 with moderate hyposmia and 9 with mild hyposmia. The associated taste disorder was hypogeusia) and 5 subjects had taste alterations only of the type hypogeusia. We did not find ageusia in any case. The 51 patients affected with chemosensory disorders correspond to 41.12% of the infected population. These data contrast with the information of the previously mentioned authors. It is possible that the type of olfactory test used in the evaluation of the cases has relevance in the results, since there are some substances that the patients cannot recognize because they do not belong to their olfactory and / or taste culture. For example, in the UPSIT test there are substances such as lilac, turpentine or peanut butter that are not identified by many of the Venezuelan patients and can sometimes be mistakenly considered pathological. Hence, in our country we designed a smell test adapted to the Venezuelan population (8,9) The other explanation for these results is related to the period of time in which this report was made, since the rate of coronavirus infections in Venezuela, it began to show exponential growth since May of this year.

We did not find significant differences in terms of the type of disorder, age and sex, although the predominant type of olfactory disorder was the combination of both disorders, in males. These results are similar to those observed by Mackenzie et al (14) in the systematic review of several published series on smell and taste alterations due to COVID-19.

In relation to the cases that presented smell and taste disorders and whose diagnostic tests were negative, it is difficult to determine whether the finding is related to another viral etiology or the moment in which the RT-PCR tests were performed for SARS-CoV-2 and the rapid test corresponded to a window period and warranted a second or third sample collection for follow-up and observation of seroconversion to that described in the methodology. Negative results from the RT-PCR test do not necessarily indicate that the person has not contracted an infectious disease, as other individual factors, such as risk of exposure and possible laboratory errors, must also be considered. False negative results may occur if the specimen has not been properly collected, transported, handled, or treated, does not exclude infection, and should not be the sole basis for treating patients. (15) After approximately one week from the first clinical manifestations, the sensitivity of molecular diagnosis (PCR) gradually decreases for SARS-CoV-2 infections, due to the decrease in the number of virus particles in the epithelium of the tract respiratory. In such cases, patients may have false negative results, despite ongoing infection. However, we also observed that in many patients, positive RT-PCR results persisted for more than 21 days.

Detection of SARS-CoV-2-specific serum antibodies allows for a rapid, cost-effective, and reasonably sensitive clinical diagnosis of COVID-19, as immunoglobulins such as IgM provide the initial humoral response during the early stage of viral infection, prior to initiation of the adaptive high-affinity IgG response essential for long-term immune memory. Research indicates that after SARS infection, IgM class antibodies can be detected in the patient’s blood approximately 6 days after infection, while IgG can already be detected after 8 days. Since SARS-CoV-2 belongs to the same large family of viruses, which includes those that cause Middle East Respiratory Syndrome (MERS) and Severe Acute Respiratory Syndrome (SARS), it must be assumed that the process of producing antibodies it will be similar to that of other viruses belonging to this family, while the detection of IgG antibodies and IgM antibodies that act against SARS-CoV-2 may be an indication of infection. Furthermore, the detection of IgM antibodies generally indicates a recent exposure to SARS-CoV-2, whereas the detection of IgG antibodies in the case of COVID-19 indicates exposure to the virus some time ago (16)

When using tests that detect IgM and IgG antibodies, it should be remembered that a positive result may be evidence of a past infection, not an active infection. Negative results of serological tests do not exclude SARS-CoV-2 infection, as the ‘window period’ (delay in the production of antibodies) can exceed 7 days. Serological tests can also give false positive results. This may be the case of a past or ongoing infection with virus strains other than SARS-CoV-2, such as the HKU1, NL63, OC43, or 229E coronavirus.

Therefore, serological tests are applied as a complementary method to monitor the epidemiological situation, although they can be performed faster and are less expensive than genetic tests. This diagnostic method has limited sensitivity, but efforts to improve it are ongoing as it is useful in controlling the infection. Due to insufficient data on, inter alia, the dynamics of the immune response to infection and the diagnostic value of the tests available to detect IgM and IgG class antibodies (including sensitivity, specificity, positive and negative predictive value), In many countries, the use of serologic tests for diagnostic purposes is currently not recommended. (16) With our findings regarding patients who were symptomatic and with smell and taste disorders but negative by RT-PCR for SARS-CoV-2 With a positive rapid test and the situation in which they presented negative results by SARS-CoV-2 RT-PCR but with chemosensory disorders, we concluded two situations: at the time of evaluation they did not present sufficient antibody titers for diagnosis and later they could seroconvert or b. Not all symptomatic patients are COVID-19, nor should all individuals with chemosensory disorders necessarily be classified as infected with coronavirus.

Recently Brann et al. (2020) have published the possible mechanisms by which olfactory and taste alterations can occur in patients with SARS-CoV-2 infection and it has been advocated that these represent the best predictor factor for coronavirus infection, however the Positive predictive value of chemosensory disorders in our population does not coincide with these statements. The sensitivity shown was 39.29% and specificity of 67.23% of the chemosensory test in the diagnosis of COVID-19. The positive Predictive Value of the smell and taste test is 57.3% and the Negative Predictive Value is 54.09%. This means that the probability that an individual with a positive RT-PCR test for SARS-CoV-2 and a chemosensory test is positive is 57%. These findings also do not agree with the Moein-Vaira series, where chemosensory disorder is considered. as a predictor of disease.

18.29% of symptomatic patients reported chemosensory disorders before doing the RT-PCR test for SARS-CoV-2, most of them manifested it during their hospitalization and it was corroborated with the test (see table 5). These findings do not coincide with large series such as those described by Mackenzie et al (2020) where chemosensory alterations are the best predictors of coronavirus infection. The proportion of patients with general symptoms such as fever, headache, myalgias, arthralgias, chills, hyporexia, general malaise, not associated with smell and taste disorders (31 cases), is similar to the patients who have these symptoms associated with the disorder (see table 5) in contrast to the large series reported by Mackenzie et al.

Regarding the recovery time for smell and taste alterations, Hopkins (2020) reports that they are transitory and the recovery time varies between 5 days and 4 weeks, depending on the severity of the symptoms. (17) These findings they were carried out through patient surveys which are subjective and whose follow-up is doubtful. In this investigation, there were 12 patients with anosmia (2 as the only manifestation of the disease). In the 124 positive patients for SARS-CoV-2, we were able to carry out a complete follow-up with smell and taste tests and RT-PCR from the beginning, corroborating their spontaneous recovery in the vast majority of them. Only one patient persists with anosmia-type smell disorder, and in her other types of evaluations were carried out to establish the real cause of it. At this point there is a similarity with what is reported in the literature, in terms of recovery time. In our review of other investigations, we did not find any relationship between the persistence of chemosensory alterations and the negativization of RT-PCR. There appears to be no relationship between improvement in the disorder and viral load detected by molecular and / or serological tests.

The low positive predictive value between the positivity of the chemosensory disorder and the coronavirus infection or the presence of 42 asymptomatic patients positive for SARS-CoV-2 in our study, could be due to several factors previously described, such as the time in which the test began. study, early at the peak of infections, the lack of performance of 2 or more PCR-RT SARS-CoV-2 tests to follow-up suspected cases or hypothetical factors among which the race, aspects related to the type of genetic mutations of the virus and migratory movement. The mutation of the SARS-CoV-2 spike, which mediates infection in human cells, has been described. Korber et al (2020) have specified 13 mutations, which are considered in a broader phylogenetic spectrum and can change with geography and over time, which may confer some selective advantages in transmission or resistance to infection of some populations. In the case of Europe, the Spike (spike) D614G mutation was the dominant one and began to spread in February 2020, causing such aggressive behavior in the population. There is evidence of recombination of strains that circulate locally and that are indicative of infections by multiple strains (18) This aspect is currently being investigated in Colombia and Venezuela (18) Rodríguez-Morales et al. (18) described 3 genomic sequences of coronavirus mutations that can explain the behavior of the disease in Venezuela, in specific areas such as Zulia, (in which a significant migratory movement of people to and from Colombia and Brazil has been observed) where the severity of the infection has been greater than those of other geographical areas of the country (18) This hypothesis could provide an explanation to understand why infections are mild or asymptomatic in some areas, severe in others, or why the penetrance of chemosensory disorders is lower than in other reports.

## CONCLUSIONS AND SUGGESTIONS

In this research we cannot consider smell and taste disorders as predictive factors of SARS-CoV-2 disease, but rather as clinical manifestations that patients may have during the coronavirus infection and that are associated with other symptoms The patients evolve spontaneously and recover in a period between 3 days to 5 weeks with an average of 8.5 days. We only observed one patient who had not recovered her sense of smell until the end of this research phase, which corresponds to 120 days. More than 30% of the positive population is asymptomatic, so it is recommended to carry out SARS-CoV-2 RT-PCR on a large scale, and / or the implementation of the smell and taste test that allow screening of a larger number of people, for diagnosis and control of disease progression. It is recommended to expand the sample of future research to corroborate the findings presented in this study.

## THANKS

Very special thanks to all those people who contributed in some way in the processing of samples, analysis, data collection and anteroom before the direction of hospitals and services involved. Special mention to Dr. José Manuel García, Dr. Francisco Sánchez, Dr. Isabel Duque, Dr. David Forero, Ptte (Ej) Margarett Tovar, Ptte (Ej) María Barico.

10 Health Centers participated in the study and were the following:

University Military Hospital “Dr. Carlos Arvelo “(San Martín-Caracas), Hospital” Dr. José Ignacio Baldó “, (Algodonal-Caracas) General Hospital” Dr. Jesús Yerena “(Lídice-Caracas), Ophthalmological Hospital” Dr. Francisco Risquez “(Cotiza-Caracas), Hospital de los Seguros Sociales” Dr. Miguel Pérez Carreño “(Caracas), Hospital of Social Security” Dr. José María Vargas “(La Guaira),” Nelson Sayago Mora “Military Hospital (La Asunción-Nueva Esparta),” Dr.Vicente Salias “Military Hospital (Fuerte Tiuna-Caracas) Metropolitan Polyclinic (Miranda State) University Clinical Hospital of Caracas. Other cases screening centers by the General Health Directorate of the national armed forces located in Greater Caracas: General Command of the Army and Aviation, Headquarters of Seguros Horizonte, Headquarters of the Military Prosecutor’s Office, Headquarters of TVFANB, Headquarters of the Bank of the National Armed Forces. Some commercial premises located in Miranda State that wish to remain anonymous also participated in the collection of data and patient evaluation, as well as house-to-house registration.

## Data Availability

all data can be used

https://doi.org/10.1101/2020.04.29.069054

https://www.medrxiv.org/content/10.1101/2020.08.25.20182063v1

## FINANCING

The resources for the rapid tests and the virocult used, as well as other implements for sampling and processing, were provided by the Ministry of Popular Power for Health of Venezuela.

The disposable smell test kits were developed and self-financed by its authors, Dres Rodriguez Wilneg, Velazquez Carlos, and Pieruzzini Rosalinda.

The taste test was carried out with samples of the 5 basic flavors

## CONFLICTS OF INTEREST

The authors of this research do not declare conflicts of interest.

## Notes

### Competing Interest Statement

The authors have declared no competing interest.

### Funding Statement

Self-financed by the authors and the Ministry of People's Power for Health

### Author Declarations

The Bioethics Commission of the Hospital Militar Universitario Dr. Carlos Arvelo

## BIBLIOGRAPHY

1. Lechien J,Chiesa-Estoma C,De Siati D,Horoi M,Le Bon S et al.Olfactory and gustatory dysfunctions as a clinical presentation of mild to moderate forms of the coronavirus disease (COVID-19).A Multicenter European Study.European Archives of Oto-Rhino-Laryngology.Published online April 6 2020.

2. Gane S,Kelly C,Hopkins C. (2020)Isolated Suddenonset anosmia in COVID-19 infection.A novel Syndrome?Special Report.Open Acces.Rhinology 58:0,0-0.https://doi.org/10,4193/Rhin20.114.

3. Giacomella A, Pezzati L,Conti F,Bernacchia D,Siano M,Oreni L,Rusconi S,Gervasoni C,Ridolfo A,Rizzardini G,Antinori S,Galli M.Self-reported Olfactory and Taste Disorders in patients with severe acute respiratory coronavirus 2 infection: A cross-sectional study.Letter to Editor.Clinical infectious Diseases. https://doi.org/10.1093/cid/ciaa330.

4. Ramanathan K,Antognini D,Combes A,Paden M,Zakahry B,Ogino M,MacLaren G,Shekar K (2020)Planning and provision of ECMO services for severe ARDS during the COVID-19 pandemicand other outbreaks of emerging infectious diseases.Lancet respir Med.2213-2600(20)30121–1.

5. Wu YC,Chen Cs,Chan YJ (2020) Overview of the novel coronavirus (2019nCov):the patogen of severe specific contagious pneumonia (SSCP):J Chin Med Assoc. https://doi.org/10.1097/JCMA000000000000000270

6. Brann D,Tsukahara T,Weinreb C,Logan D,Data S.(2020) Non Neural expression of SARS-CoV-2 entry genes in the olfactory epithelium suggest mechanisms underlying anosmia in COVID-19 patients. Science Advanced, vol. 6, no. 31, eabc5801 DOI: 10.1126/sciadv.abc5801

7. Netland J,Meyerholz DK,Morre S, Cassell M,Perlman S. Severe Acute respiratory síndrome coronavirus infection causes neuronal death in the absenceof encephalitis in mice transgenic for human ACE2. J.Virol.2008;82 (15):7264–7275. doi:10.1128/JVI.00737-08.

8. Pieruzzini R,Rodríguez W, Velazquez C. Short Adaptation of the smell test of the University of Pennsylvania (UPSIT) for the Venezuelan population.International Archives of Otorhynolaryngology.2020.In press.

9. Hagobian A, Pieruzzini R. Hospimil Smell Test vs Connecticut Test for the Diagnosis of Olfactory Disorders. Presented at the X Triológico Venezolano de Otorrinolaringología, November 11-13, 2019, for publication in Acta Otorrinolaringológica. 2020.

10. Castro L, Pieruzzini R, Luque E, Garcia L. Proposal of a Taste Test for the Venezuelan population. Otorhinolaryngology Act.Vol 31 nº 01 2020.Pp 17–26.

11. Spinato G,Fabbris C,Polesel J,Cazzador D,Borsetto D,Hopkins C,Boscolo-R P,(2020)Alteratiosn in Smell or taste in midly Symtomatic Outpatients with SARS-CoV-2 Infection.JAMA.https://jamanetwork.com

12. Viera LA,Deiana G,Fois Giuseppe A,Pirina P,Maddedu G, De Vito A, et al.Objective Evaluation of anosmia and ageusia in patients with COVID-19:experience in unique center with 72 cases.(2020)Head and neck.DOI:10.1002/hed.26204.

13. Moein S, Hashemian S, Mansourafshar B, Khorram ‐ Tousi A, Tabarsi P, and Doty R.Olfactory Dysfunction.A biomarker of COVID-19.Int Forum Alergia Rhinol. 2020 17 de abril: 10.1002/alr.22587.

14. Mackenzie E, Ramirez V, Lipson S, Herriman R, Toskala A, Lin C, Joseph P, Reed D.Objective sensory testing methods revealing a higher prevalence of olfactory loss in patients with COVID-19 in comparative to subjective methods: a sistematic review and methanalysis. Versión 1. medRxiv. Preimpresión 2020 6 july.

15. Kubina R, Dziedzic A.Molecular and Serological Tests for COVID-19. A Comparative Review of SARS-CoV-2 CoronavirusLaboratory and Point-of-Care Diagnostics. Diagnostics (Basel) 2020 Jun; 10(6): 434. Published online 2020 Jun 26. doi: 10.3390/diagnostics10060434.

16. Li Z., Yi Y., Luo X., Xiong N., Liu Y., Li S., Sun R., Wang Y., Hu B., Chen W., et al.Development and clinical application of a rapid IgM-IgG combined test for SARS-CoV-2 infection diagnosis. J Med Virol. 2020 27 de febrero; 10.1002/jmv.25727.doi: 10.1002/jmv.25727.

17. Hopkins C,Surda P,Whitehead E,Kumar B.Early recovery following new onset anosmia during the Covid-19 pandemic.An observational cohort study.J.Otolaryngology Head and neck surgery. (2020);May 4:49:26

17. Korber B,Fisher WM,Gnanakaran S,Yoon H,Theiler J,Abfalterer W,Foley B,Giorgi EE,Bhattacharya T,Parker M,Partridge DG,Evans CM,freeman TM,De silva T.Spike mutation pipeline reveals the emergency of more transmisible of SARS-CoV-2.bioRxiv 2020.04.29.doi: https://doi.org/10.1101/2020.04.29.069054

18. Rodriguez-Morales A,Balbin-Ramon G,Rabaan A,Sah r,dhama K,Paniz-Mondolfi Genomic Epidemiology and its importance in the study of the COVID-19 pandemic.Le infezioni in Meicina,nº2,139–142,2020.

